# Estimation of salt intake, potassium intake and sodium-to-potassium ratio by 24-hour urinary excretion: an urban rural study in Sri Lanka

**DOI:** 10.1101/2020.04.17.20068833

**Authors:** R Jayatissa, Y Yamori, AH De Silva, M Mori, PC De Silva, KH De Silva

**Affiliations:** Department of Nutrition, Medical Research Institute, Colombo; Mickagowa University, Japan; Ministry of Health, Colombo

**Keywords:** urine sodium, urine potassium, salt, blood pressure, sodium-potassium-ratio

## Abstract

**Background:** Sodium intakes of different populations around the world became of interest after a positive correlation was drawn between dietary sodium intake and prevalence of hypertension. Sri Lanka has adopted a salt reduction strategy to combat high blood pressure in the population with escalation of non-communicable diseases.

**Objective:** To measure intake of salt, potassium and sodium/potassium ratio of adults in urban and rural settings.

**Design:** A community based study of 328 adults between 30-59 years, including equal numbers from urban and rural sectors. Weight, height and waist circumference were measured. Blood pressure was measured by a standardized automated measurement system and the mean of two readings was used for analysis. 24-hour urine was collected and measured for creatinine, sodium, potassium levels.

**Results:** Mean daily salt consumption was 8.3g (95%CI:7.9,8.8), which is 1.6 times higher than WHO recommendation. Mean daily potassium intake was 1,265g (95%CI:1191.0,1339.3), which is 2.8 times lower and sodium/potassium ratio was 4.3 (95%CI:4.2,4.5), which is 7 times higher than WHO recommendation. Daily salt consumption was significantly higher in males (9.0g;95%CI:8.3,9.8) than females (7.7g;95%CI:7.2,8.2); rural (8.9g;95%CI:8.2-9.6,) than urban (7.7g;95%CI:7.2,8.3) with increasing body mass index (8.2g;95%CI:6.1,10.2 to 10.0g;95%CI:8.5,11.6). Systolic blood pressure was significantly positively correlated with high BMI and waist circumference.

**Conclusions:** High salt consumption, low potassium intake and high sodium/potassium ratio was found in this population. This information can be used to set targets to reduce salt intake in the population. Need to create awareness to enhance the consumption of potassium rich food while reducing salt intake to minimize future NCD burden.

## Introduction

Chronic Non Communicable Diseases (NCDs), particularly Cardio Vascular Diseases (CVD) and its principal risk factor High Blood Pressure (HBP), are increasing in the South East Asia Region (SEAR)^1^. It was found leading cause of CVD were attributed to hypertension in the South East Asia Region^2^. High sodium intake has been identified as a major risk factor for hypertension^1^. Reducing the population’s sodium intake has been recommended as an effective and cost effective intervention to improve health outcome at population level^3^.

World Health Organization recommends a salt intake of less than 5 grams (approximately 2 grams sodium) per person per day and reducing salt intake by 6 g/day in populations equates to approximately 2.5 million preventable deaths globally every year^1,4^. Worldwide data on salt consumption is still limited. Considering the changing dietary patterns of today such as increasing use of processed foods and ready-made food purchased from food vendors, the estimated population salt intake in countries is much higher than the recommended target of 5g/day. Recent studies revealed that voluntary salt reductions by the food industry would save billions of health care cost^5^.

Studies have shown that high sodium intake, low potassium intake and higher ratios of sodium to potassium significantly increase the risk of CVD^3^. It further indicated that a high sodium/potassium ratio was a stronger indicator in identifying the increased risk of CVDs than levels of either sodium or potassium alone^4^. It is important to identify the consumption patterns of foods low in sodium and high in potassium of the general population as a protective measure in preventing CVDs.

In Sri Lanka, ministry of health data indicated that 71% of all annual deaths in Sri Lanka were due to chronic NCDs. Among all NCDs, 29.6% of causes of mortality, morbidity and disability accounting for CVDs^6^. Causes of CVD were hypertensive heart diseases (18.9%), ischemic heart disease (29.9%), cerebrovascular diseases (14.9%) and other causes (36.3%)^7^. The prevalence of hypertension in Sri Lankan adult men and women was 13% and 14% respectively in 2008^8^.

Daily salt intake among Sri Lankans is considered to be high and estimated as 10.5g. Major source (70-80%) is from cooked food at the level of households adding salt while cooking and the contribution from the processed food was 15-20%. The Ministry of Health (MoH) in 2010 introduced “salt reduction initiative”, which aimed at reducing daily dietary salt intake by conducting the public awareness campaign. It also aimed at reducing the salt content in all processed foods closely working with the food industry^9^. However, in order to implement the salt reduction programme in Sri Lanka, MoH needed the baseline information to set the targets to monitor intakes of salt. Measuring and monitoring population sodium/salt consumption therefore be an important step for Sri Lanka. Under this situation, measuring the sodium in 24-h excretion of urine was very useful to start programmes and to monitor salt levels^,9^. Hence this study was aimed to determine the salt intake, potassium intake and sodium/potassium ratio in urban and rural populations in Sri Lanka.

## Methodology

It was a cross sectional community survey. Study population was healthy adults (both male and female) between the aged 30-59 years. Calculated sample size was 360 to ensure the possibility to detect difference between urban/rural and considering 95% precision, 1.5 design effect and 10% non-response rate. Ethical clearance was obtained from the ethics committee of Medical Research Institute (MRI), Colombo. The sample was drawn from the mostly populated province, which was divided into 2 strata (urban and rural). Then 20 public health midwife (PHM) areas, which is the smallest health area consists of approximately 3000 population and 600 households, were randomly selected (10 from each strata) based on probability proportional to size sampling. Each PHM area was divided into 12 blocks, each with 50 households using the map considering the 50% of the population belonged to the adults 30-59 years. One block was randomly selected for the study. First households to be visited was randomly selected. Then nearest households to the right was visited next and one individual per household was randomly selected to participate in the survey, equal numbers from male/female and each age group (30-39, 40-49 and 50-59) to fulfil 18 from each PHM area. All the selected adults were asked to attend the closet health center on a predetermined date. Weight, height and waist circumference were checked according to standard protocols by the trained MRI staff^10^. Blood pressure was measured by a standardized automated measurement system after allowing participant to sit for 5 minutes by the medical officer. The mean of 2 readings was used for analysis^11^.

Survey staff explained how to collect 24-hour urine and written instructions were provided. Participants were instructed to discard the first morning void and to collect all urine over the following 24-hour, including the first void on the next morning and to record the time of start and finish collection. Each participant was provided with the aliquot cup, explanation sheet and a disposable 5 liter capacity plastic bag to collect urine^11^. The plastic bag was especially designed for portability to ensure collection if respondents could not be home for some of the collection period. After completing 24-hour urine collection, all participants brought back the bag filled with urine to the health center on the same day.

All urine samples were labeled and dispatched to the MRI laboratory and weighed. Then a sample of urine was frozen without preservatives at −20°C and send to the laboratory of Mickagowa University, Japan for estimation of urinary sodium (mEq/day), urinary potassium (mEq/day) and urinary creatinine (mg/day) using indirect ion-selective electrodes methodology. 24 hour urine output was measured by deducting the weight of the bag which was 100g. Sodium (Na) and potassium (K) level of the urine was analysed in mEq/L and Na/K ratio was calculated as sodium divided by potassium.

### Data analysis

Body mass index (BMI) was estimated as weight in kilograms/(height in meters^2^) and categorized using WHO cut-off levels^10^. Central obesity was defined as waist circumference ≥ 90 cm for male and ≥ 80 cm for female. Hypertension was defined with the presence of systolic blood pressure (SBP) ≥ 130 mmHg or diastolic blood pressure (DBP) ≥ 85 mmHg. Completeness of the 24-hour urine collection was assessed calculating the creatinine coefficient (CC), which is creatinine excretion (mg/day) / bodyweight in kg. Creatinine coefficient of 14.4 to 33.6 in men and 10.8 to 25.2 in women were considered as an acceptable 24-hour urine collection^11^. Sodium level was multiplied by 24 hour urine volume to calculate 24 hour sodium levels. Daily salt intake was estimated based on calculation of 24-hour urinary sodium excretion assuming all sodium ingested was in the form of sodium chloride. Salt intake in grams was calculated considering 1mEq of sodium was equal to 23mg and 1g NaCl was equal to 393.4mg of sodium^11^. Daily salt intake was divided into quintiles as follows: <5.5, 5.5-7.6, 7.7-10.6 to >10.6 g/day. Mean salt intake and 95% confidence levels (95%CI) in relation to urban/rural, male/female and age groups were calculated. Simple Pearson correlation coefficients (r) was used to identify and test the strength of a relationship between urinary sodium, potassium, sodium-to-potassium ratio with systolic and diastolic blood pressure levels. A p-value <0.05 was considered to indicate statistical significance. All data were analyzed by using SPSS, version 23.

## Results

A total of 328 adults between 30-59 years; 165 from urban and 163 from rural sector, were included in the study. Response rate variation from total to urban to rural sector was 91.1%, 91.6% and 90.6% respectively. Creatinine coefficient was 17.6 in men and 12.9 in women. The basic characteristics of the study population in urban/rural sectors are shown in Table 1. There is no significant difference between age and sex distribution of the study population in both sectors. Proportion of overweight and obesity in the urban participants (40.6% and 13.9%) was significantly higher than the rural participants (29.4% and 5.5%). Mean age, SBP and DBP was not significantly different between two sectors. Mean BMI and mean WC was significantly higher in the urban counterparts (25.2±4.4kg/m^2^; 89.8±11.1cm) than the rural counterparts (23.9 ±3.9kg/m^2^; 86.6±8.7cm).

**Table 1:** Basic characteristics of the study population.

| Basic Characteristics | No. (%) / mean (SD) |  | Total |
| --- | --- | --- | --- |
|  | Urban | Rural |  |
| Age group (years) |  |  |  |
| 30-39 | 26.7 | 31.9 | 106 (30.7) |
| 40-49 | 35.2 | 34.4 | 116 (33.6) |
| 50-59 | 38.2 | 33.7 | 123 (35.7) |
| Mean age (years) | 45.1 (8.4) | 44.8 (8.2) | 45.2 (8.3) |
| Sex |  |  |  |
| Male | 47.3 | 45.4 | 152 (46.3) |
| Female | 52.7 | 54.6 | 176 (53.7) |
| BMI Status (kg/m<sup>2</sup>)*** |  |  |  |
| <18.5 (underweight) | 6.1 | 5.7 | 21 (6.4) |
| 18.5-24.9 (normal) | 39.4 | 57.7 | 159 (48.5) |
| 25.0-29.9 (overweight) | 40.6 | 30.1 | 116 (35.4) |
| >=30 (obesity) | 13.9 | 5.5 | 32 (9.8) |
| Mean BMI (kg/m<sup>2</sup>)*** | 25.2 (4.4) | 23.9 (3.8) | 24.5 (4.1) |
| Mean waist circumference (cm)** | 89.0 (11.4) | 86.3 (9.0) | 87.6 (10.4) |
| Mean systolic BP (mmHg) | 130.2(18.1) | 127.6(17.3) | 128.9(17.7) |
| Mean diastolic BP (mmHg) | 76.9 (12.4) | 74.3(11.2) | 75.6 (11.9) |
| Total | 165 (50.3) | 163 (49.7) | 328 |

\*\*, \*\*\*indicates significance at the 5% and 1% level, respectively

As shown in Table 2, there is no difference of 24-hour urine volume between urban and rural sectors, average was 1.27±0.7 liter/day. Mean creatinine coefficient was significantly difference between urban and rural subjects (13.2Vs16.9). Mean urinary sodium and potassium was significantly higher in rural counterparts than urban (3,223.9Vs2,788.9mg/day) and (1,374.5Vs1,161.5 mg/day) respectively. Of all study subjects, 30.5% had sodium intake above the WHO recommendation of 2,000 mg/day, which was significantly higher in urban than rural counterparts (37.6%Vs23.3%), while only 0.6% showed potassium intake above the WHO recommendation of 3,510 mg/day.

**Table 2:**
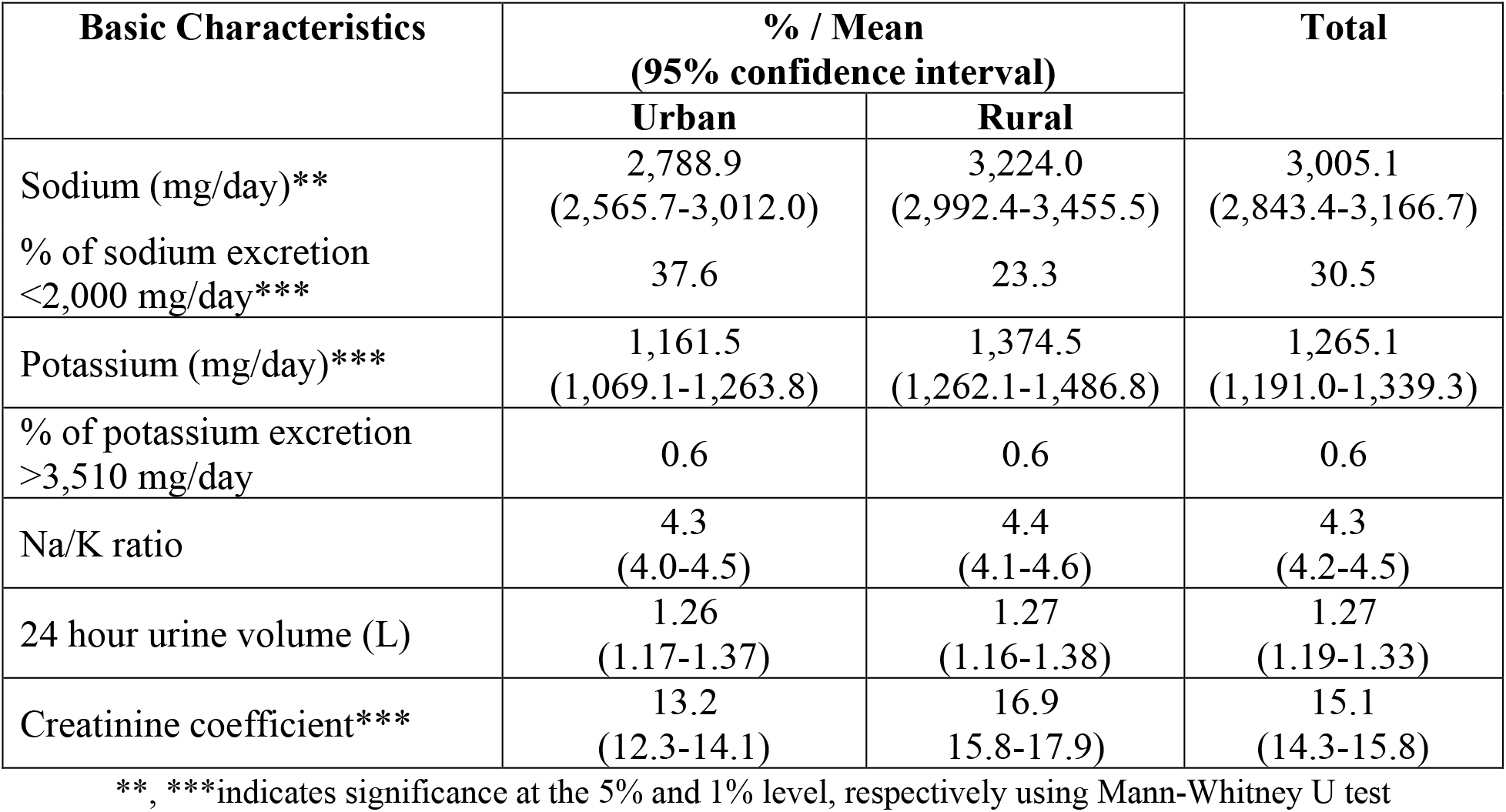
Urinary excretions of the study population in different sectors (n=328)

| Basic Characteristics | % / Mean<br>(95% confidence interval) |  | Total |
| --- | --- | --- | --- |
|  | Urban | Rural |  |
| Sodium (mg/day)** | 2,788.9<br>(2,565.7-3,012.0) | 3,224.0<br>(2,992.4-3,455.5) | 3,005.1<br>(2,843.4-3,166.7) |
| % of sodium excretion<br><2,000 mg/day*** | 37.6 | 23.3 | 30.5 |
| Potassium (mg/day)*** | 1,161.5<br>(1,069.1-1,263.8) | 1,374.5<br>(1,262.1-1,486.8) | 1,265.1<br>(1,191.0-1,339.3) |
| % of potassium excretion<br>>3,510 mg/day | 0.6 | 0.6 | 0.6 |
| Na/K ratio | 4.3<br>(4.0-4.5) | 4.4<br>(4.1-4.6) | 4.3<br>(4.2-4.5) |
| 24 hour urine volume (L) | 1.26<br>(1.17-1.37) | 1.27<br>(1.16-1.38) | 1.27<br>(1.19-1.33) |
| Creatinine coefficient*** | 13.2<br>(12.3-14.1) | 16.9<br>(15.8-17.9) | 15.1<br>(14.3-15.8) |

\*\*, \*\*\*indicates significance at the 5% and 1% level, respectively using Mann-Whitney U test

As presented in Table 3, mean daily salt (NaCl) consumption of the study population was 8.3g (95%CI:7.9,8.8g) and Na/K ratio was 4.3 (95%CI:4.2,4.5). Daily salt consumption was significantly higher in males than females (9.0Vs7.7g); rural than urban (8.9Vs7.7g); obese subjects than overweight, normal and underweight subjects (10.0Vs8.7, 7.7 and 8.2g respectively). However, mean salt consumption did not significantly differ with presence or absence of central obesity as well as between hypertensives and non-hypertensives and between age groups. Mean Na/K ratio was significantly decreasing with the increasing age from 4.7 to 4.0.

**Table 3:** Mean daily consumption of the salt (NaCl) in grams and sodium-to-potassium ratio in relation to basic characteristics of the study population.

| Basic Characteristics | Mean consumption (95% confidence interval) |  |  |
| --- | --- | --- | --- |
|  | No. | Salt | Na/K ratio |
| **Age group (years)** |  | *p=0.5* | *p=0.02* |
| 30-39 | 96 | 8.5 (7.9-9.2) | 4.7 (4.3-5.0) |
| 40-49 | 114 | 8.2 (7.4-9.0) | 4.4 (4.1-4.7) |
| 50-59 | 118 | 8.2 (7.4-9.1) | 4.0 (3.8-4.3) |
| **Sex** |  | *p=0.004* | *p=0.12* |
| Male | 152 | 9.0 (8.3-9.8) | 4.5 (4.2-4.7) |
| Female | 176 | 7.7 (7.2-8.2) | 4.2 (4.0-4.5) |
| **Sector** |  | *p=0.02* | *p=1.0* |
| Urban | 165 | 7.7 (7.2-8.3) | 4.4 (4.1-4.6) |
| Rural | 163 | 8.9 (8.2-9.6) | 4.4 (4.1-4.6) |
| **BMI Status (kg/m<sup>2</sup>)** |  | *p=0.005* | *p=0.6* |
| <18.5 (underweight) | 21 | 8.2 (6.1-10.2) | 4.6 (3.6-5.7) |
| 18.5-24.9 (normal) | 159 | 7.7 (7.1-8.4) | 4.3 (4.0-4.6) |
| 25.0-29.9 (overweight) | 116 | 8.7 (8.0-9.4) | 4.3 (4.0-4.6) |
| >=30 (obesity) | 32 | 10.0 (8.5-11.6) | 4.6 (4.2-5.1) |
| **Hypertension** |  | *p=0.9* | *p=0.7* |
| Yes | 132 | 8.5 (7.7-9.3) | 4.3 (4.0-4.5) |
| No | 196 | 8.2 (7.7-8.7) | 4.4 (4.1-4.6) |
| **Central obesity** |  | *p=0.6* | *p=0.3* |
| Yes | 207 | 8.3 (7.8-8.8) | 4.3 (4.1-4.5) |
| No | 121 | 8.3 (7.5-9.2) | 4.4 (4.1-4.8) |
|  | **328** | **8.3 (7.9-8.8)** | **4.3 (4.2-4.5)** |

p values by Mann-Whitney U test

Table 4 shows the daily consumption of salt quintiles in study subjects. One third of female (30.1%) were in the lowest quintile, in contrast one third of male (30.9%) were in the highest quintile. Study subjects with highest mean BMI (25.8±4.8kg/m^2^) and highest mean WC (91.6±10.3cm) were in the highest quintile group. Both these observations were statistically significant. No significant difference between salt quintiles and mean age, systolic and diastolic BP.

**Table 4:** Characteristics of the study population by the daily consumption of salt quintiles.

| Characteristics | Daily salt consumption in grams (% or Mean± SD) |  |  |  | P value |
| --- | --- | --- | --- | --- | --- |
|  | <5.5 | 5.5-7.6 | 7.7-10.6 | >10.6 |  |
| **Sex** |  |  |  |  |  |
| Male | 18.4 | 24.3 | 26.3 | 30.9 |  |
| Female | 30.1 | 26.1 | 23.9 | 19.9 | **P=0.03** |
| **Sector** |  |  |  |  |  |
| Urban | 28.5 | 23.0 | 28.5 | 20.0 |  |
| Rural | 20.9 | 27.6 | 21.5 | 30.1 | P=0.06 |
| Age (years) | 46.3 ± 8.0 | 44.5 ± 7.7 | 45.3 ± 8.8 | 44.7 ± 8.6 | P=0.5 |
| BMI (kg/m<sup>2</sup>) | 24.0 ± 3.8 | 23.7 ± 3.9 | 24.8 ± 3.9 | 25.8 ± 4.8 | **P=0.005** |
| Systolic BP (mm Hg) | 127.4 ± 18.1 | 125.1 ± 15.9 | 126.9±17.1 | 124.6 ± 16.9 | P=0.7 |
| Diastolic BP (mm Hg) | 75.4 ± 12.6 | 74.3 ± 11.5 | 73.9±11.6 | 74.7 ± 11.4 | P=0.9 |
| Waist circumference (cm) | 86.5 ± 10.6 | 84.4 ± 9.9 | 88.1±9.6 | 91.6 ± 10.3 | **P=0.000** |

Table 5 shows that age, BMI, WC, being a male were significant positively correlated with systolic and diastolic BP. In contrast urinary sodium, potassium, sodium intake and sodium-to-potassium ratio did not show a similar correlation.

**Table 3:**
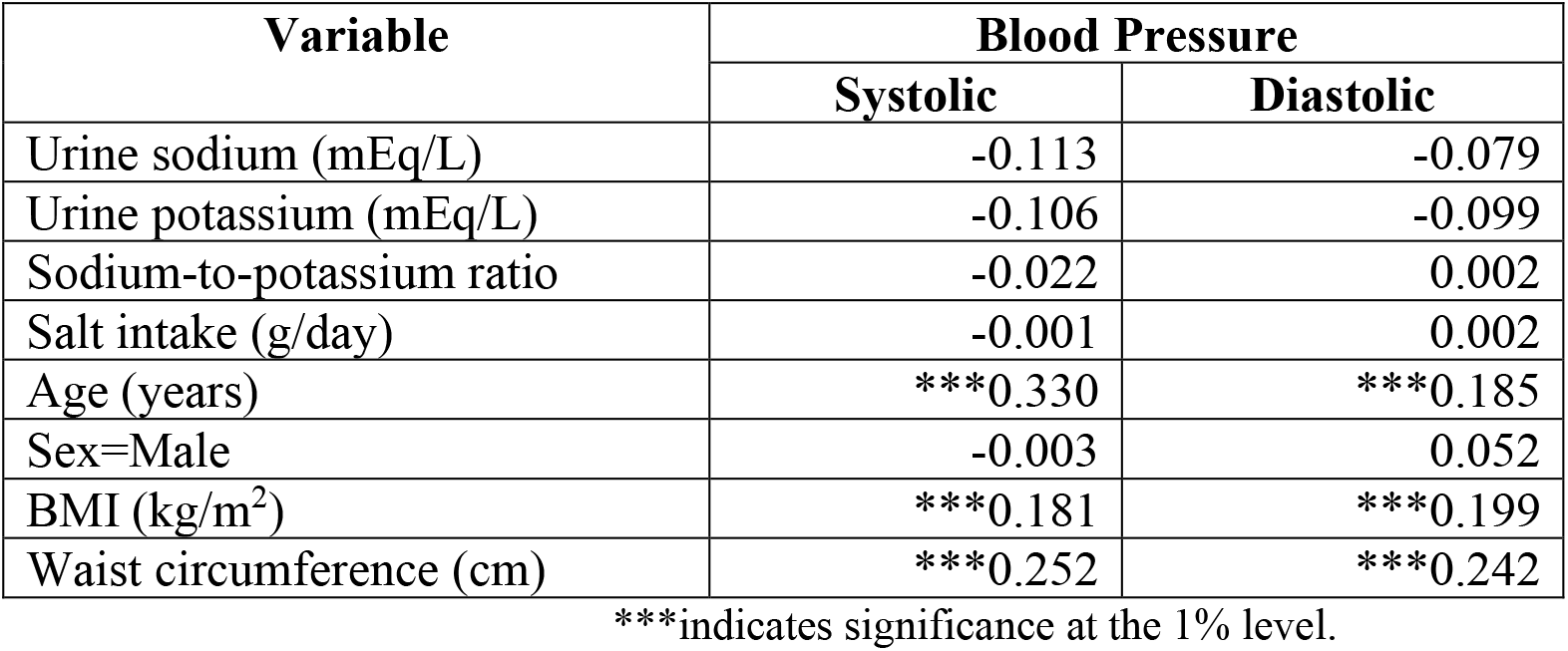
Correlations Coefficient (r) between blood pressure and 24hr urinary sodium, potassium, sodium/potassium ratio and other factors (N=298)

## Discussion

Our study is the first in Sri Lanka to estimate sodium and potassium intake in a representative sample of adults aged 30-59 years using 24-hour urine excretion. This study estimated an overall daily intake of salt as 8.3g, which is 1.6 times higher than the WHO recommendation of 5g/day. In 1970, dietary data indicated daily salt intake was 7g in Sri Lanka^12^. Comparing our results with studies in other countries, similar results were found in India (8.6g)^13^, Australia (9.0g)^14^, UK (8.1g)^15^ and higher levels in China (11.8g)^16^.

The amount of daily sodium excretion in our study was 3005.1mg (95%CI:2843.4,3166.7), which is 1.5 times higher than the WHO recommendation of 2000mg/day^11^. It was observed significantly higher sodium excretion in rural participants (2788.9mg) compared to urban (3223.9mg). There is a significantly higher daily salt intake in rural participants (8.9g) than urban (7.7g). However, participants in the urban and rural areas showed similar daily volume of urine (1.26LVs1.27L respectively) indicating the difference in cooking practices between urban and rural sector. In Sri Lanka, majority (>70%) consumed home cooked food. Our results have shown that 69.5% of participants exceed the WHO recommended sodium intake^11^. Other reported studies showed the similar findings^13,14^.

The amount of daily potassium excretion in our study was 1265.1mg(95%CI:1191.0,1339.3), which is 2.8 times lower than the WHO recommendation of 3,510mg/day^11^. Significantly higher potassium excretion in rural participants than urban (1161.2mgVs1370.3mg respectively). Our results have shown that 99.4% of participants did not meet WHO recommended potassium intake^11^. It indicates the low consumption of potassium rich food among participants. Results of other studies are in line with our findings^12,13,14,15^.

Na/K ratio is an important risk factor for hypertension (WHO 2007). In our study mean urinary Na/K ratio was 4.3, while 4.3 in urban and 4.4 in rural sectors^17,18^. However, the ratio in our study subjects is seven times greater than the WHO recommended 0.59, which indicates the poor eating behavior of study subjects.

Our study also examined the association of urban/rural, age, sex, BMI categories, central obesity, hypertension with daily salt intake and Na/K ratio. This study showed a significant relationship between body mass index, sector and waist circumference with salt intake. This indicates indirectly that the type of foods consumed by overweight and obese people eat more quantities of food or more salty food than the persons who are underweight and with normal BMI. In the follow-up to the first National Health and Nutrition Examination Survey, He et al^19^ reported a 61% increase in cardiovascular disease mortality in overweight persons and 39% increase in all-cause mortality associated with 2.3g of higher sodium intake. Across the board, sodium excretion was greater in men than women, probably due to their higher food intake. For instance, a survey carried out in UK, has observed an intake of 10g of salt/day in men and 7.7g in women^20^.

This study further showed higher quintile of daily salt consumption is associated with men, high BMI and high WC highlighting to focus on these groups for salt reduction interventions. Our study showed the urinary sodium, potassium, salt intake and Na/K ratio are not related to the level of systolic and diastolic blood pressure, this may be due to not controlling confounding factors. As shown in this study, it appears that minor changes in BMI and WC may result in a decrease in the occurrence of high blood pressure. This indicates the importance of consuming foods high in potassium in parallel to the reduction of sodium in the diet^21,22,23,24^. Further research to identify the association between sodium and BP at Sri Lankan settings is warranted^25^.

A strength of our study is using 24-hour urine excretion, which is the gold standard for assessing daily salt intake in a population as it captures > 90% of the sodium ingested within a day^13^. High participating rate of 91.1% indicates good population estimates in this study^14^. Creatinine coefficient was 17.6 in men and 12.9 in women indicated an acceptable 24-hour urine collection of this sample^15^. Limitation of this study was not considering confounding factors such as physical activity, energy intake during performing linear regression. The location of the study being limited to only one province of Sri Lanka compromises the direct generalizability of the study findings to Sri Lanka as a whole. However, this is the only study which is available to obtain baseline information required to assess the impact of the salt reduction strategy.

## Conclusions

Our study found Sri Lankan adults have high sodium intake and low potassium intake compared to the WHO recommendations. It is recommended to create awareness among the population about the effects on high salt intake especially among overweight / obese people to minimize health consequences while increasing the potassium rich food. It is recommended to implement of an effective salt reduction programme as a priority programme in Sri Lanka.

## Data Availability

Available with corresponding author

## Acknowledgements

We would like to thank health staff of selected health areas for assisting us in conducting the study. Staff of the Department of Nutrition, Medical Research Institute in collecting data and samples.

## Author Contributions

Renuka Jayatissa (RJ), Y. Yamori (YY), A.H. De Silva (AHS), Mari Mori (MM) were responsible for the study concept and study design and laboratory analysis of data. RJ was responsible for data acquisition and performing statistical analysis. Both RJ and PC De Silva (PCS) equally contributed to preparation of manuscript and data analysis.

